# Changes in the prevalence of antimalarial partner drug resistance markers and policy in six sub-Saharan African countries from 2000 to 2021: A systematic review

**DOI:** 10.1101/2025.05.08.25327217

**Authors:** Alexandra Walker, Amanda Ross, Christian Nsanzabana

## Abstract

**Background:** Prompt malaria case management is a cornerstone for malaria control and elimination. However, this strategy is threatened by the development of antimalarial drug resistance. Resistance is mediated through spontaneous genetic changes such as mutations, but drug pressure is the main driver of resistance spread. Molecular markers of resistance may provide insight into spatiotemporal dynamics of drug resistance, and how drug policy changes may affect the spread of resistance.

**Methods:** We conducted a systematic review to assess the dynamics of *Pfcrt, Pfmdr1, Pfdhfr* and *Pfdhps* mutations from 2000 to 2021. Six countries from sub-Saharan Africa were selected based on availability of molecular data, and varying antimalarial drug policies; Kenya, Malawi and Uganda in East Africa and Burkina Faso, Cote d’Ivoire and Nigeria in West Africa. Medline, Embase, Cochrane and Elsevier databases were searched for relevant literature and identified records were screened for prevalence data and extracted.

**Results:** Overall, 138 studies were included. Estimated prevalence of *Pfcrt* 76T declined following cessation of CQ, though at variable levels between countries. All countries saw an increase in *Pfmdr1* N86/D1246 prevalence, with faster increases in East Africa, while *Pfmdr1* 184F prevalence increased, except in Burkina Faso. Prevalence of *Pfdhfr* (51I/59R/108N) and *Pfdhps* (436A/437G/540E) mutations reached fixation levels in most countries, however, the 164L and 581G mutations, increased during the time period only in Kenya and Uganda.

**Conclusions:** Our study provides compelling evidence on the impact of antimalarial drug policy change on molecular markers of resistance, and their potential use to monitor drug resistance spread.

## Background

Over the last 20 years, a major change in antimalarial drug policy has occurred worldwide, with countries shifting towards the use of artemisinin based combination therapies (ACTs) (1). A pivotal factor in shaping global antimalarial drug policy was the emergence of drug resistance to the previously used drugs, chloroquine (CQ) and sulfadoxine-pyrimethamine (SP) (2). Although efficacy of ACTs for the treatment of uncomplicated *Plasmodium falciparum* infection remains high in sub-Saharan Africa, drug resistance is a threat to both the artemisinin compound and the partner drug (3).

Partial resistance to artemisinin was first identified in Cambodia in 2008 (4) and subsequently in other areas of the Greater Mekong Subregion (GMS) (5, 6). The ensuing failure of ACTs to clear the infection and a lack of novel therapeutics has made treatment in the GMS difficult (7). Recently, independent emergence of artemisinin resistance has been reported in a number of East African countries (8-10). As the majority of malaria cases and deaths occur in sub-Saharan Africa, where transmission remains high in many settings (11), a situation similar to the GMS could have disastrous consequences (12). It is therefore vital that the spread of antimalarial drug resistance to both artemisinin and partner drugs is monitored and contained as much as possible.

As the frequency of exposure to a drug increases within a community, so does the drug pressure, driving selection and leading to the further spread of resistant parasite clones (13). However, the impact of policy changes, such as changes in first-line treatment or preventative chemotherapies such as intermittent preventative treatment in pregnancy (IPTp) and Seasonal Malaria Chemoprevention (SMC), is not well-established.

The use of surveillance tools to observe spatial and temporal trends can give policy makers an idea of past, present and future trends of drug resistance. One strategy is to track molecular markers associated with antimalarial drug resistance (14). Mutations involving *P. falciparum chloroquine resistance transporter* (*Pfcrt)* and *P. falciparum multidrug resistance protein 1* (*Pfmdr1)* genes are commonly assessed for ACT partner drug resistance, as the presence of either the wild type or mutant clone is associated with reduced susceptibility to lumefantrine or amodiaquine respectively (15). As SP is still commonly used in preventative chemotherapies, mutations in *P. falciparum dihydropteroate synthetase* (*Pfdhps*) and *P. falciparum dihydrofolate reductase* (*Pfdhfr*) are associated with sulfadoxine and pyrimethamine resistance respectively and are commonly assessed in conjunction (16). The prevalence levels and changing dynamics of these markers over time may be influenced by antimalarial drug policy changes. By understanding how these policies influence molecular markers of drug resistance, there is the potential to inform future policies.

In this systematic review, we aimed to describe the changes in the prevalence of molecular markers associated with ACT partner drugs and preventative chemotherapies occurring after the introduction or cessation of key drug-based interventions or drug policy changes. As East and West Africa have historically used contrasting policies over different periods, these two regions were selected for this study, with three countries in each region selected to represent regional diversity.

## Methods

The Preferred Reporting Items for Systematic Reviews and Meta-Analysis (PRISMA) statement was used as a framework to guide the conduct and reporting of this systematic review (17). The outcome measures selected were the prevalence of *Pfcrt, Pfmdr1, Pfdhfr* and *Pfdhps* gene polymorphisms in each country, between 2000 and 2021. Molecular markers that represent the currently used partner drugs in ACTs or drugs used in preventative chemotherapies and have previously been validated according to the WHO were chosen (18) (Table 1).

**Table 1.**
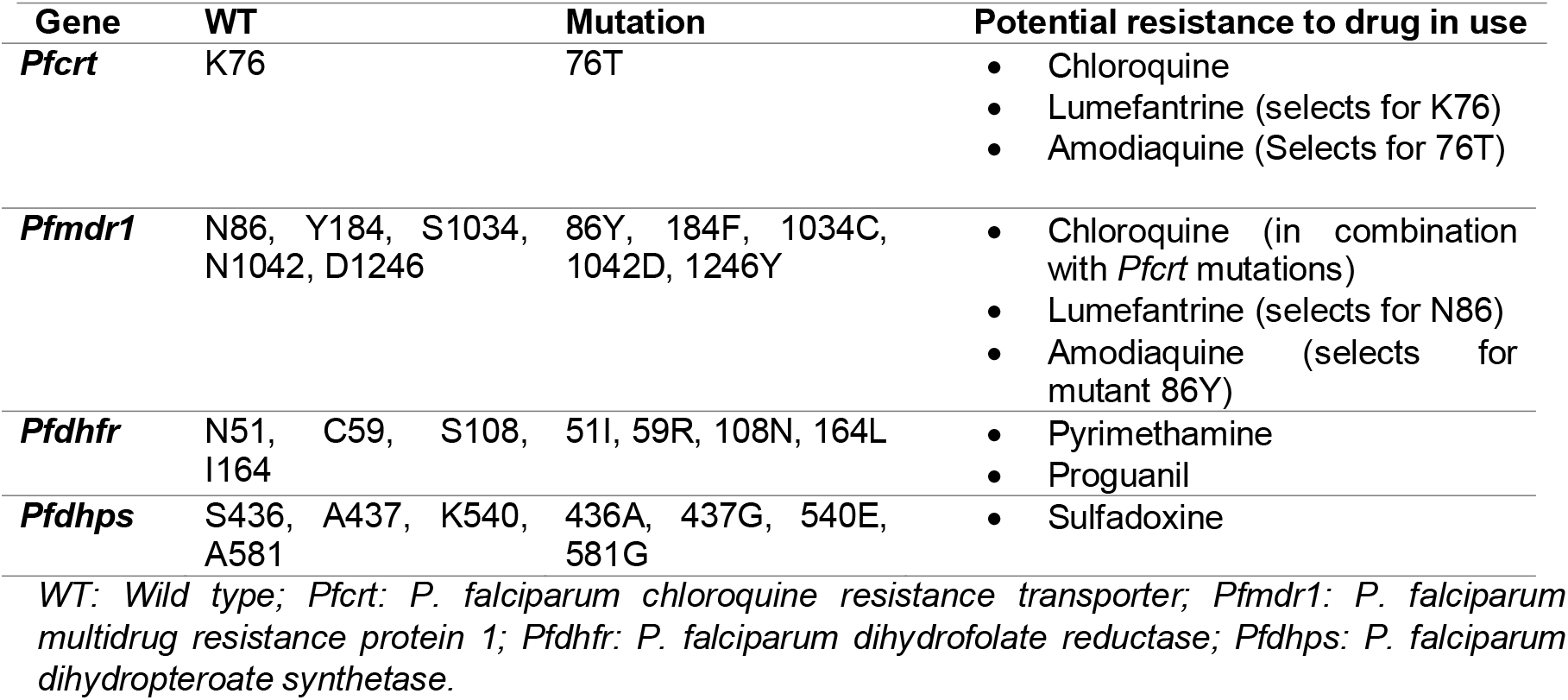
Molecular markers associated with resistance to antimalarial drugs included in this review. The table is adapted from (18).

Countries were selected with the aim that each would provide a unique profile of the prevalence of drug resistance markers, based on their historical and current antimalarial drug use. In total six countries were selected: Kenya, Malawi and Uganda in East Africa, and Burkina Faso, Cote d’Ivoire and Nigeria were selected for West Africa.

### Search strategy

Four electronic databases were searched: PubMed, Embase, Scopus and Cochrane. An initial search was performed in October 2022, encompassing literature from 1 January 2000 to 18 October 2022. The search was updated in September 2023 to include any additional studies that collected data in the time period. Controlled vocabularies, synonyms and indexed terms were included when possible. Full search terms are available in Supplementary information 1, Table S1. The Worldwide Antimalarial Resistance Network (WWARN) molecular surveyor database was also searched for additional records or unpublished data (19).

### Study selection and screening

Citations for identified records were imported into Rayyan (20) directly or using Endnote software (20). Following identification, confirmation and removal of duplicates, one individual performed title and abstract screening, followed by full text screening according to the eligibility criteria (Supplementary information 1, Table S2). The criteria included original research within a selected country, during the selected time period and prevalence of one or more of the selected markers of antimalarial drug resistance.

### Data extraction

Data from eligible publications were then extracted, including country of origin, year of collection, the number of genotyped samples and the number of samples carrying each molecular marker of interest. Additional data collected included study design, age group, genotyping method and district of collection.

### Data analysis

Study characteristics were summarized using frequencies and percentages, per country. The prevalence of associated mutations for each study was plotted over time (2000 to 2021) for each country and gene separately. The timing of changes in antimalarial drug policy for the recommended first line treatment of uncomplicated *P. falciparum* treatment and preventative chemotherapies were marked on the plot. This allows the patterns in prevalence to be visualised relative to the timing of policy changes. Point size was proportional to the number of samples that were genotyped for each study. If a study reported both single and mixed genotypes of a mutation, these were combined to give the total number of positive samples. For prevalence data collected over multiple years (equal to or less than three years), the data was combined into a single time point, represented by the midpoint of the collection period

All analyses were conducted in R (Version 4.3.2) (21).

## Results

### Study characteristics

A search of the literature returned 1,409 publications (Figure 1). Of these, 629 duplicate records were removed before screening and 551 were excluded based on the selection criteria. A total of 229 records were sought for retrieval, with an additional 22 identified during extraction.

**Figure 1.**
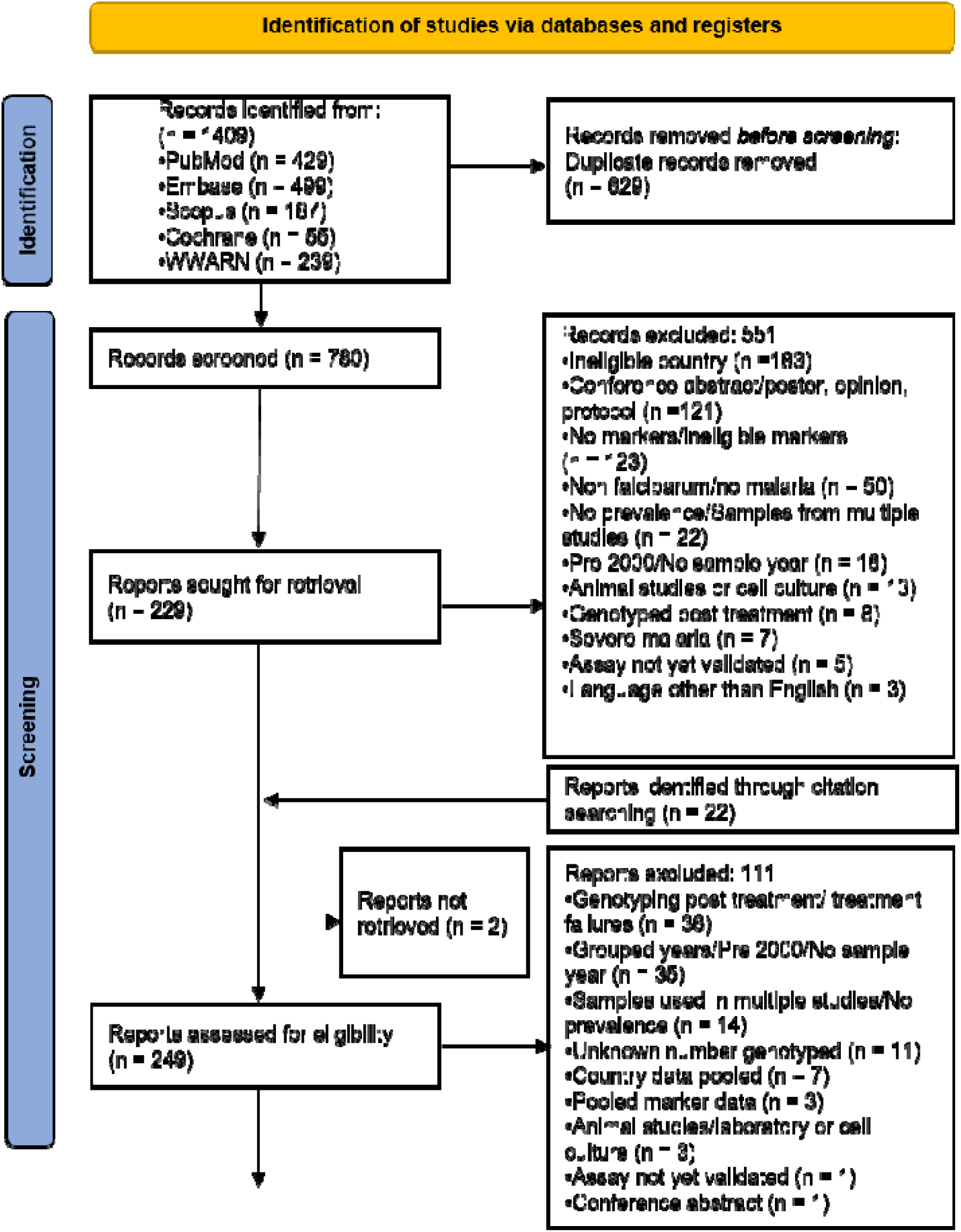
PRISMA flowchart for the identification and selection of studies

Overall, of the 249 full reports retrieved, 111 failed to meet the selection criteria leaving a total of 138 studies eligible for inclusion. Sixty-nine of the studies were from the selected East African countries, fifty-eight studies from West African countries and eleven from multi-country studies (Supplementary information 2, Table S1). No formal risk of bias was performed during extraction, though risk of bias was assessed for the pattern over time rather than for individual studies.

The years of sample collection for the included studies were spread between 2000 and 2021 (Figure 2), with the highest frequencies between 2010 and 2018. Collection of samples occurred throughout the time-period for most countries, with the exception of Malawi which had no studies included after 2012 and Cote d’Ivoire which had a low overall number of included studies and intermittent collection years. Collection for multi-country studies, of which imported cases were included, primarily occurred after 2010.

**Figure 2.**
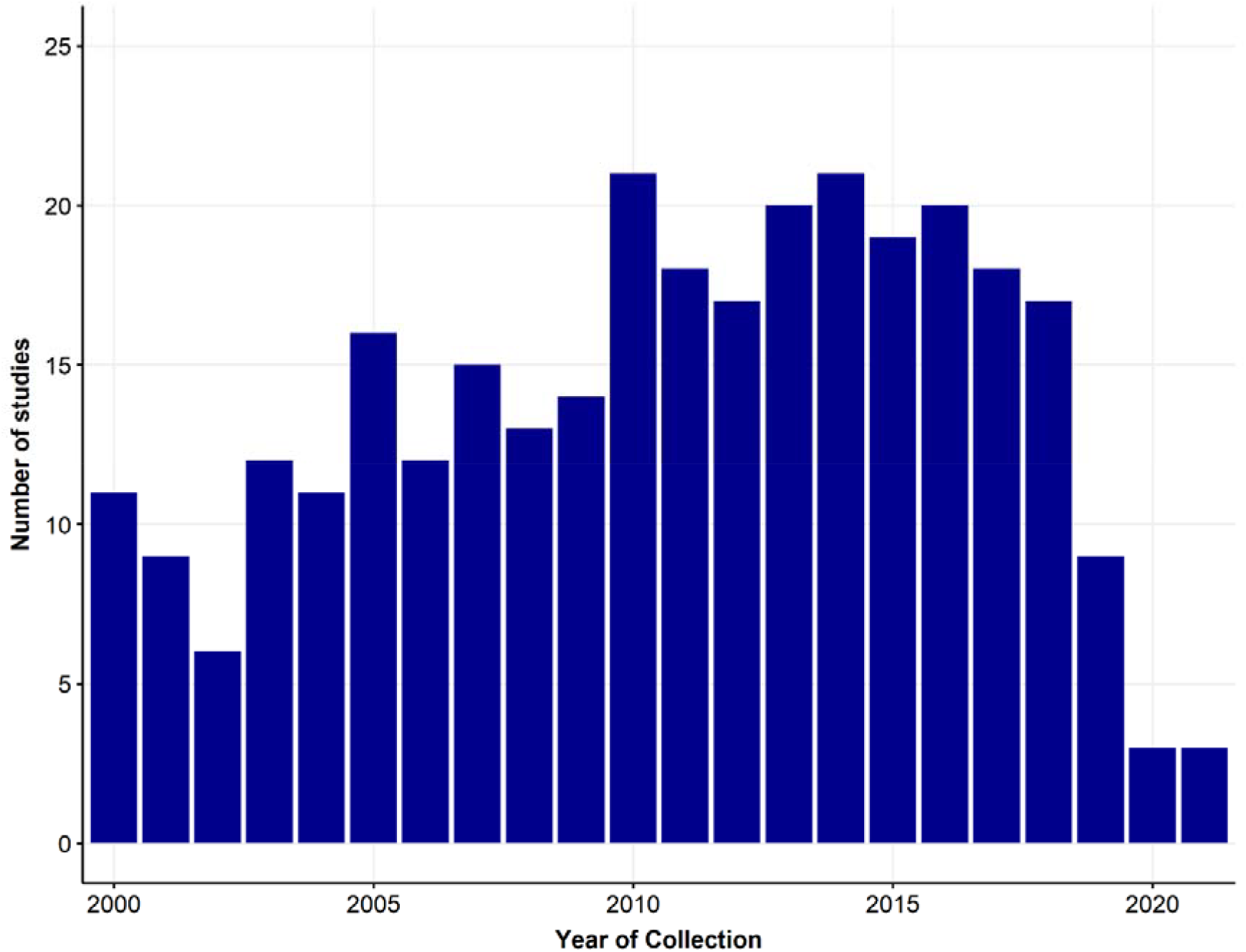
Study Data Collection Frequency per Year (Studies with multiple collection years appear once for each year in the dataset)

Overall, *Pfcrt* was the gene genotyped in the highest number of studies (n = 88/138, 64%) and in each country, with the exception of Kenya and Nigeria where *Pfmdr1* was the most genotyped gene (Table 2). Few studies genotyped all genes (n = 24/138, 17%).

**Table 2.**
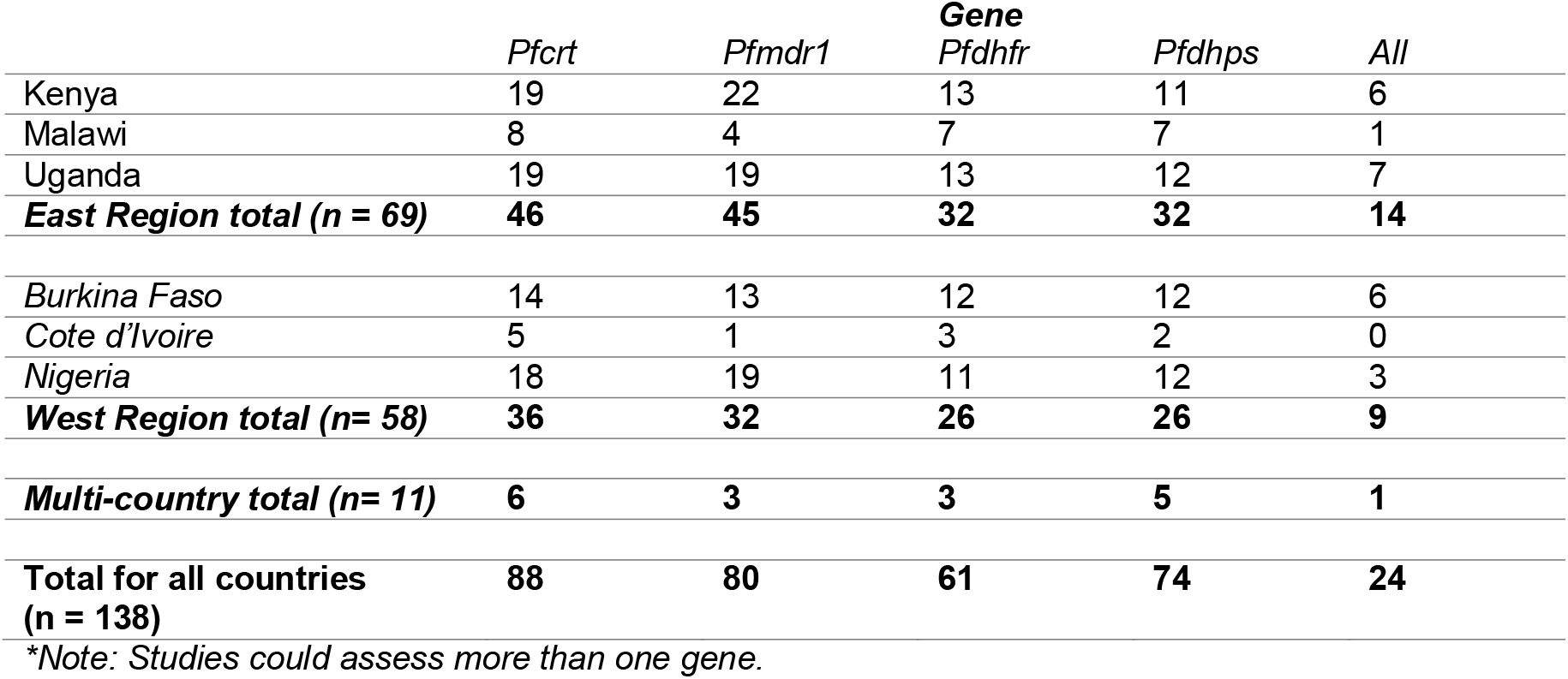
Number of studies for each marker by region and country.

Approximately, half of the studies used a cross-sectional study design (n = 76/138, 55%), and Restriction Fragment Length Polymorphism (RFLP) and PCR/sequencing were the dominant methods for genotyping (Supplementary information 2, Table S2). Children under 18 years old were the most frequently sampled age group, with more than one-third of studies sampling children between the ages of 3 and 59 months (n = 25/65, 38%) (Supplementary information 2, Table S2). Collection varied between districts in each country, with sample collection often heavily concentrated in specific areas (Supplementary information 2, Figure S1 and S2).

### Estimated prevalence of *Pfcrt* 76T mutation

Between 2000 and 2021, all countries included in this study experienced a decline in *Pfcrt* 76T mutation prevalence, following the first-line treatment change away from CQ (Figure 3). In West Africa, in the years after the cessation of CQ, 76T mutations continued to rise, before a declining trend was observed (Figure 3), even though, from 2011, the prevalence appears to remain sizeable in all three countries, demonstrating incomplete elimination of the 76T mutation. In East Africa, the prevalence of 76T remained at high levels in Kenya and Uganda up to ten years after formal cessation of CQ, before declining to negligible levels. In Malawi, 76T mutation levels were very low (< 10%) from 2000, in respect to the other countries. Overall, despite CQ cessation and a decline in 76T mutation prevalence, the mutation remained present in Burkina Faso and Nigeria in West Africa and Uganda in East Africa, between 2017 and 2021. Regional differences in each country are seen in Supplementary information 2, Figure S3.

**Figure 3.**
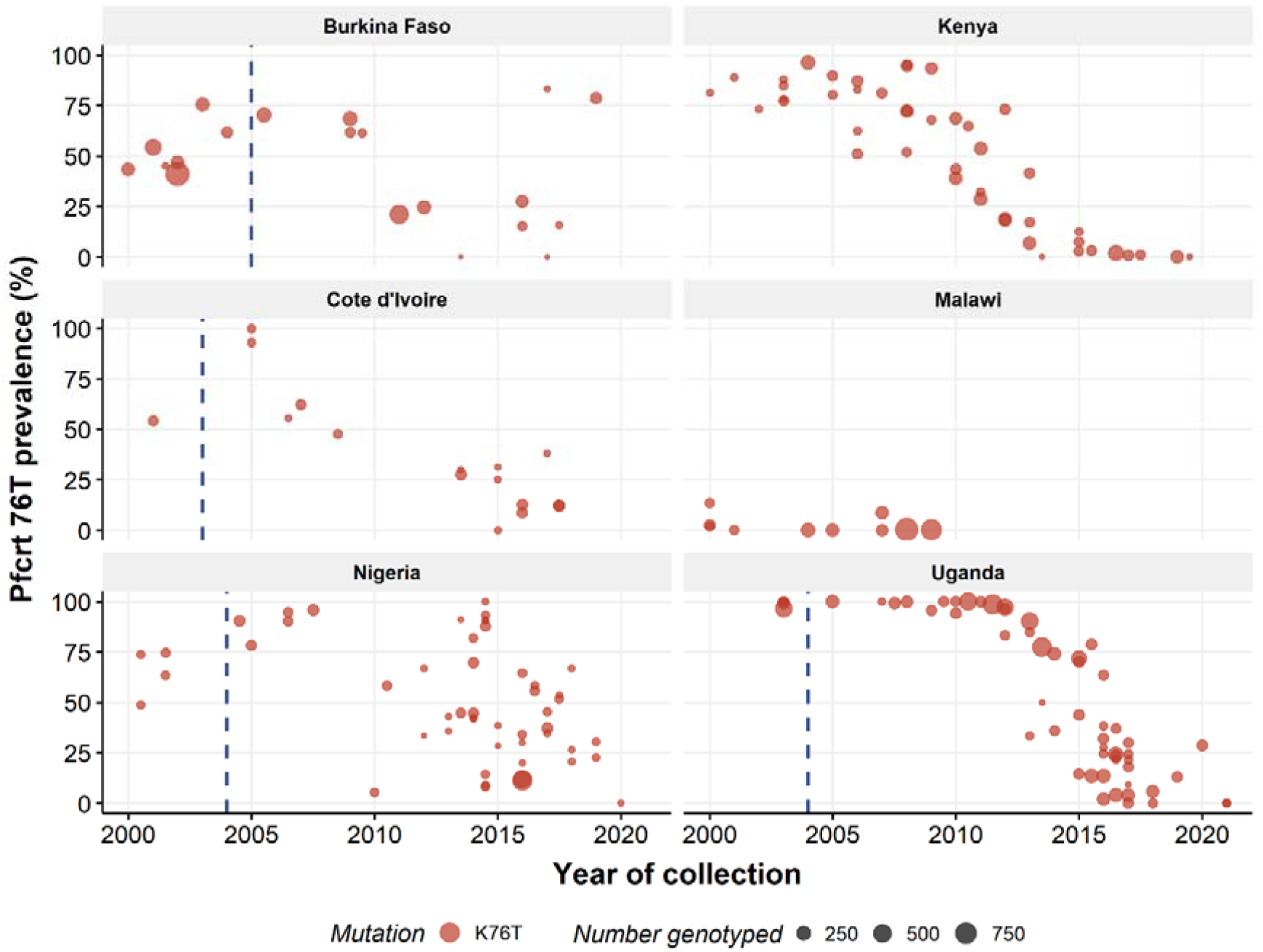
Prevalence of Pfcrt 76T mutation by country. Dashed blue line in each graph represents the cessation of CQ, with the exception of Kenya and Malawi (Burkina Faso: 2005, Cote d’Ivoire: 2004, Nigeria: 2004, Kenya: 1998, Malawi: 1993, Uganda: 2004), dashed pink line represents the initiation of SMC (Burkina Faso: 2014, Nigeria: 2013). In Uganda, CQ was used in conjunction with SP from 2000 to 2004, until cessation. Circle size is proportional to the number of samples genotyped from each study.

### Estimated prevalence of *Pfmdr1* mutations

We excluded Cote d’Ivoire, Malawi and the *Pfmdr1* 1034C and 1042D mutations since limited data was available. After the introduction of both artemether-lumefantrine (AL) and artesunate-amodiaquine (AS+AQ) to Burkina Faso in 2005, a downward trend was observed in all mutations of the *Pfmdr1* gene (Figure 4).

**Figure 4.**
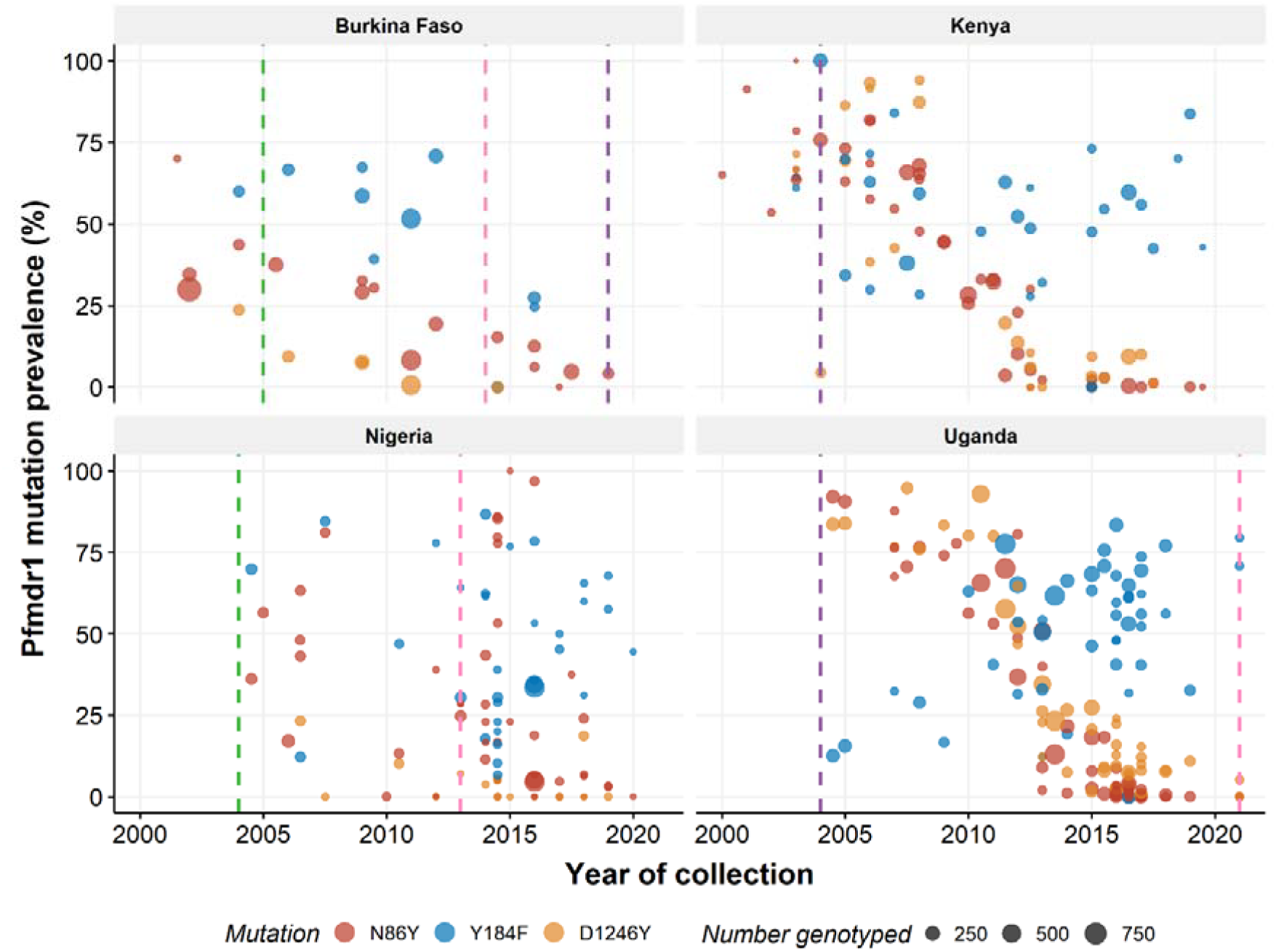
Prevalence of Pfmdr1 N86Y, Y184F and D1246Y mutations by country. Policy change lines are included for chosen first-line antimalarial drugs affecting mutations, including both AS+AQ and AL (green dashed line), AL only (purple dashed line) and the initiation of SMC (pink dashed line). Circle size is proportional to the number of samples genotyped from each study. Colours correspond to the mutation that is present.

However, the same trends were not observed for Nigeria, who also introduced both AL and AS+AQ in 2004, with the prevalence of 86Y and 184F varying greatly within a year, and over time (86Y: 0-86% in 2014 and 0-96% in 2016, 184F: 6-87% in 2014). Although a number of studies were conducted post-SMC rollout (2013), there is no clear trend, though there is within-region heterogeneity present as well as between region variation (Supplementary information 2, Figure S4).

In contrast, Kenya and Uganda in East Africa introduced AL as the only recommended first-line antimalarial treatment from 2004 and in both countries a substantial decrease was observed for 86Y and 1246Y after this time. Before 2004 in Kenya, the prevalence of all *Pfmdr1* mutations was above 50%, with the subsequent years showing a slight increase before decreasing throughout the time period. The estimated prevalence of the 184F mutation fluctuates over time but follows a general upward trend, though prevalence varies between regions (Supplementary information 2, Figure S4). Though no studies were performed before the introduction of AL, similar trends were seen in the studies from Uganda, in all regions. Both 86Y and 1246Y are shown to decline sharply from 2010, demonstrating minimal prevalence in more recent years (86Y: approximately 1% from 2018 to 2021; 1246Y: 0-10% from 2018-2021). Conversely, the prevalence of 184F increases throughout the time period, from approximately 12% in 2004 to greater than 70% in 2021.

### Estimated prevalence of *Pfdhfr* mutations

The use of SP for IPTp was introduced in the early 2000s in all countries, with the exception of Malawi, which formally introduced IPTp in 1993. The trends of the estimated prevalence of *Pfdhfr* mutations (51I, 59R, 108N, and 164L) over time in the different countries vary by mutation (Figure 5). In Burkina Faso, 51I, 59R and 108N were at low levels in 2000, before increasing steadily from 2004 onwards to more than 90% in 2016. *Pfdhfr* mutations were also present in Nigeria before the introduction of IPTp (2004) and increased to high levels in the following years, including after SMC was introduced. One study demonstrated the presence of 164L in Nigeria, imported to China, while no studies observed the mutation in Burkina Faso. There was no data available after 2007 for Cote d’Ivoire.

**Figure 5.**
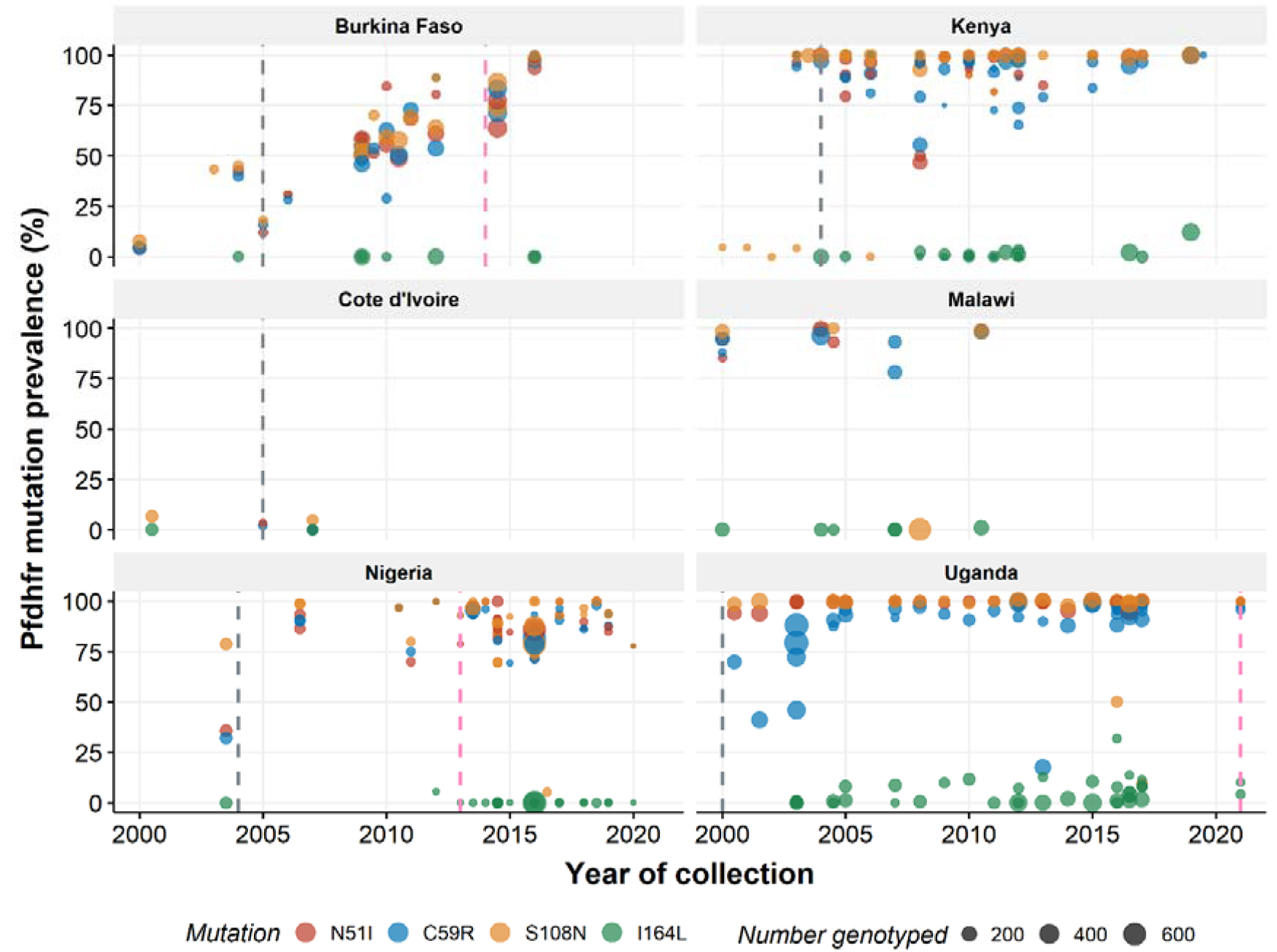
Prevalence of Pfdhfr mutations by country. Policy change lines are included for antifolate drugs used for preventative chemotherapies, including IPTp (grey dashed line) and SMC (pink dashed line). No policy line is shown for Malawi as IPTp began in 1993. Circle size is proportional to the number of samples genotyped from each study. Colours correspond to the mutation that is present.

In East Africa, the prevalence of the triple mutation (51I, 59R, 108N) was already at fixation from 2000 in each country, with the exception of one Kenyan study showing the prevalence of 108N at less than 5% between 2000 and 2006 (22). However, another study demonstrated prevalence of 51I, 59R and 108N mutations of greater than 80% in 2006, in the same region (Coast) and site (23). The 164L mutation was found intermittently at low levels throughout East Africa from 2003. In Kenya, it was only found in the Nyanza region, while in Uganda, it was present in all included regions and appeared to be increasing (Supplementary information 2, Figure S5). No data was available before the introduction of IPTp or SMC for the 164L mutation.

### Estimated prevalence of *Pfdhps* mutations

The estimated prevalence of *Pfdhps* mutations varies between East and West Africa (Figure 6). In Burkina Faso, 436A and 437G were frequently found at above 50%, and increased slightly from 2000 to 2016 while the 540E mutation was only found twice in 2003 and 2014, at a prevalence of less than 11% (24, 25). No data was available for Cote d’Ivoire after 2005. Although the majority of sample collection occurred after 2010 in Nigeria, the 437G mutation is shown to increase from 2003 as does 436A from 2005. The K540E mutation generally remained below 25% prevalence, with the exception of one study that demonstrated a prevalence of 94% in 2016 (26). The 581G mutation was also found in Nigeria. Fluctuations in mutation prevalence may be attributed to regional heterogeneity (Supplementary information 2, Figure S6).

**Figure 6.**
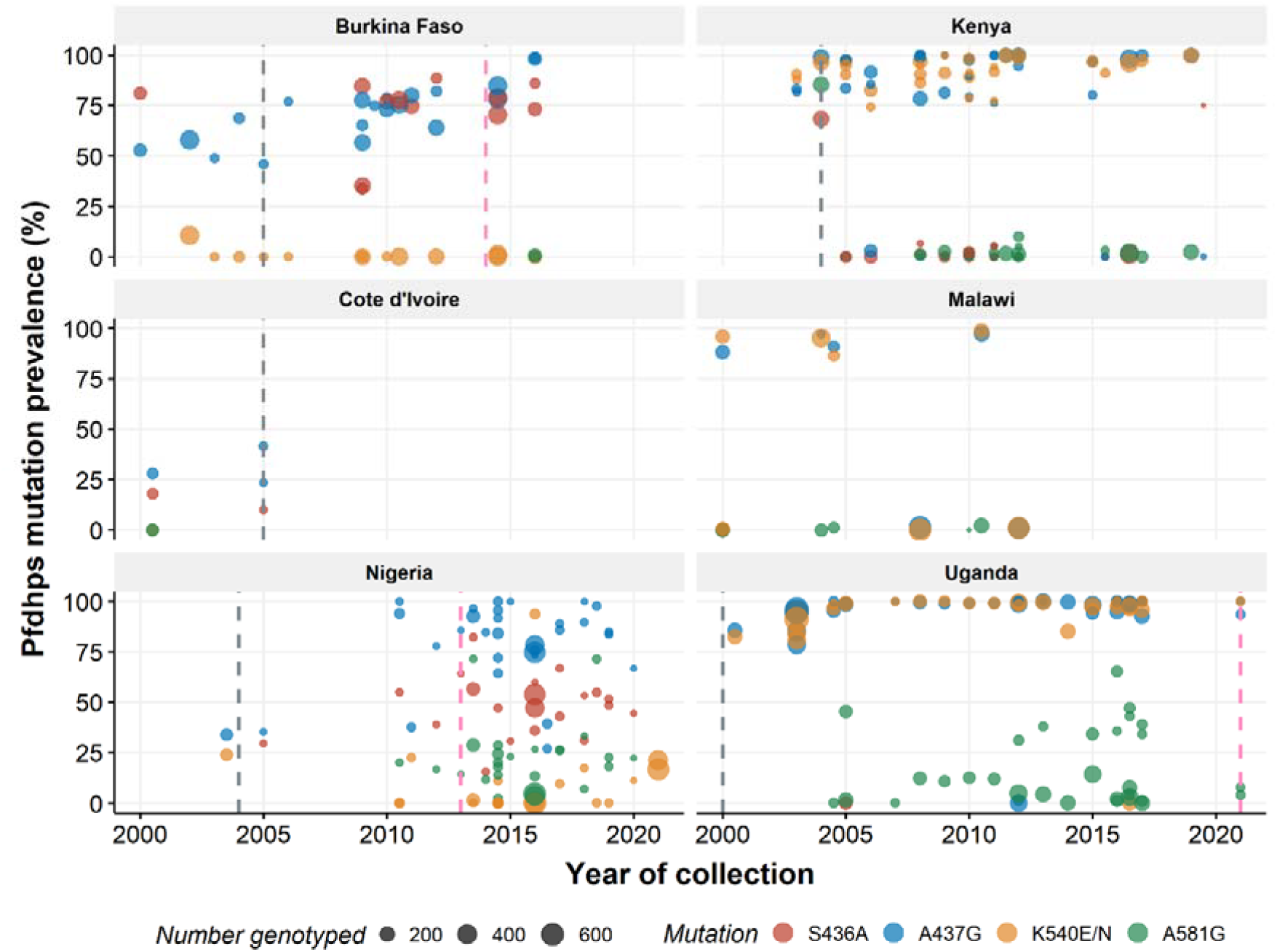
Prevalence of Pfdhps mutations by country. Policy change lines are included for antifolate drugs used for preventative chemotherapies, including IPTp (grey dashed line) and SMC (pink dashed line). No policy line is shown for Malawi as IPTp began in 1993. Circle size is proportional to the number of samples genotyped from each study. Colours correspond to the mutation that is present.

By comparison, in East Africa, 437G and 540E were the dominant mutations, primarily found at 75% or greater before or just after the initiation of IPTp and remained so throughout the time period. Although the 436A mutation was seldom genotyped and the 581G mutation was reported below 10%, a study from Kenya, conducted between 2003 and 2005, reported a 68% prevalence of the 436A mutation and 85% for the 581G mutation (27). Subsequent studies reported a maximum of 7% prevalence of 436A and 5% prevalence of 581G, within the same region and throughout the country (Supplementary information 2, Figure S6). In Uganda, the trends appear similar to Kenya, however, the 581G mutation has varied throughout the years, gradually increasing since 2004, in most regions, with a prevalence of 25% or more in the Central and Western regions (Supplementary information 2, Figure S6). The 436A mutation was genotyped once (28).

## Discussion

The increasing frequency of molecular studies since 2000 enables an updated profile on the temporal trends of antimalarial drug resistance markers. In this review, six countries were chosen from sub-Saharan Africa, with the results allowing for observations into how policy changes may have impacted the prevalence of antimalarial drug resistance markers in the past twenty years.

Cessation of CQ use as a first line treatment varied between countries (1). Consistent with previous publications, the prevalence of the *Pfcrt* 76T mutation declined in all countries, following the cessation of CQ use (22, 29-32). However, return to the K76 wild type occurred at different rates in both West and East Africa and coincided with the timing of the policy change and the region. Additional factors that may have contributed to the rate of wild type return include inherent differences among the countries such as initial prevalence, transmission intensity, access to new first line treatments, coverage levels, the availability of CQ and its continued use (32).

Following the recommendation from the WHO for the use of ACTs to treat malaria, countries within sub-Saharan Africa varied in their selection for first line treatments. The use of AS+AQ was primarily confined to West Africa, where it was used as a first line treatment with AL, while countries in East Africa, tended to use only AL. No data was collected for Central Africa in this review; however, presence of mutations has been shown to be geographically linked (33, 34), with similarities also reflected not only in the policy changes, but in the preference for antimalarial drugs (1, 35).

Although AS+AQ and AL have opposing selection pressures (15, 36, 37), the associated mutations in *Pfcrt* and *Pfmdr1* genes, showed increasing prevalence of wild type clones (K76 and N86/D1246), following the initiation of ACTs in all countries. Return to wild type *Pfmdr1* genotypes occurred faster in East Africa when compared to West Africa. This is in line with a systematic review by Okell et al. (2018) that found prevalence of 86Y and 1246Y mutations declined faster in countries that already had high rates of *Pfmdr1* mutations and used AL as a first line treatment (38). The use of both ACTs for treatment in West Africa and SMC may account for the slower decrease of 86Y and 1246Y mutations (39). These policies may also contribute to the observable trend of decreasing 184F, while in East Africa, 184F is shown to be increasing. Due to increasing ACT use and decreasing CQ availability and use, it is difficult to assess the impact that ACTs may have on 76T mutations.

As levels of CQ treatment failure increased, it was used in combination with or replaced by SP, in many East African countries, including Malawi, Kenya and Uganda (40). Indigenous mutations in the *Pfdhfr* and *Pfdhps* genes conferring low to moderate levels of resistance emerged at multiple locations throughout Africa (41) and spread from South East Asia, like *Pfcrt* mutations before it (42, 43). High levels of both *Pfdhfr* triple (51I/59R/108N) and *Pfdhps* double (437G/540E) mutations were observed in East Africa from 2000 to 2021, despite the change in policy to ACTs between 2004 and 2007 in the included countries. In West Africa, policy changes to ACTs as a first line treatment occurred after 2004, due to later failure rates of CQ (1). The prevalence of *Pfdhfr* (51I/59R/108N) and *Pfdhps* (436A/437G) gradually increased throughout the time period although SP was only recommended for IPTp and SMC (except as a treatment for uncomplicated disease in Cote d’Ivoire between 2003 and 2005) (35). A worrying trend is the increasing prevalence of *Pfdhfr* 164L and *Pfdhps* 581G in Kenya and Uganda, as increasing mutations in either gene confers higher levels of SP resistance. However, a number of studies have continued to show the protective efficacy of SP in pregnancy, despite high levels of resistance (44-46). Continued surveillance and monitoring of the protective efficacy and prevalence of antimalarial markers is required.

This study had several limitations. Only three countries were selected per region, with each of those countries identified as potentially having sufficient temporal data, for each selected molecular marker. Additionally, data is presented for each selected country on a national level, assuming no heterogeneity within countries. As each country has different levels of transmission, access and treatment rates, the results are unlikely to be representative of all districts within a country. Furthermore, collected data was primarily from the same sentinel sites within a country and typically, the same molecular markers of resistance were assessed, creating a potential selection bias. Migration patterns within and between countries, particularly border areas was also not accounted for. Using the timing of a policy change as an indicator for a change in patterns of antimalarial drug use may not have been accurate, as policy initiation can take many years to reach all areas of the country and therefore may not be representative of a change in drug use. Both AL and AS+AQ are first line treatments in Burkina Faso and Nigeria, however, no data was available to inform the proportions of use for each drug. This is particularly important with the policy change to ACTs, due to the opposing selection pressure of the drugs. Additionally, in all countries, antimalarial drugs other than the recommended first line treatment are available in pharmacies or private facilities though the proportion of use is not known.

Despite a number of limitations, this study provides a systematic review of the timing of policy changes and the temporal patterns of the prevalence of mutations in six countries. There is the potential that it can be expanded and performed for all countries in sub-Saharan Africa, with periodic updates to fill knowledge gaps in the region. This knowledge may be able to inform future policy, by providing key indicators such as time scales and prevalence limits for drug resistance markers of certain antimalarial drugs. The research may also be able to inform parameterisation of mathematical models of malaria for predicting the impact of future policy changes. Further data collection is required to accurately assess the impact of historical and current drug use on the prevalence and temporal trends of molecular markers associated with antimalarial drug resistance.

## Supporting information

PRISMA abstract

PRISMA

## Data Availability

The data that supports these findings are openly available in Zenodo at 10.5281/zenodo.15364194

https://zenodo.org/records/15364194

## Grant numbers

None

## Data availability

The datasets and code that support these findings are openly available in Zenodo at 10.5281/zenodo.15364194

This review was originally conceived as a student thesis project. It was not registered and the protocol was not published in advance.

