## Supplementary material 1 for "Changes in the prevalence of antimalarial partner drug resistance markers and policy in six sub-Saharan African countries from 2000 to 2021: A systematic review"

**Supplementary information 1**

**Search terms**

An initial search was created in PubMed, with key terms in the form of text words (both free text and title/abstract), controlled vocabularies (Medicine Medical Subject Headings (MeSH)) and indexed terms (supplementary concepts and pharmacological action) as well as spelling variations and synonyms. For other databases, the Polyglot Search Translator was used to convert the initial search blocks including controlled vocabularies and indexed terms to the relevant database (1).

*Table S1. PubMed search terms*

| #1 | ("Malaria, Falciparum"[Mesh] OR "Plasmodium falciparum"[Mesh] OR "Malaria, Falciparum"[Title/Abstract] OR "Plasmodium falciparum"[Title/Abstract]) OR malaria OR plasmodium falciparum) |
| --- | --- |
| #2 | "Burkina Faso"[MeSH Terms:noexp] OR "Cote d'Ivoire"[MeSH Terms:noexp] OR "Kenya"[MeSH Terms:noexp] OR "Malawi"[MeSH Terms:noexp] OR "Nigeria"[MeSH Terms:noexp] OR "Uganda"[MeSH Terms:noexp] OR "Burkina Faso"[All Fields] OR "Cote d'Ivoire"[All Fields] OR ("Kenya"[MeSH Terms] OR "Kenya"[All Fields] OR "kenya s"[All Fields]) OR ("Malawi"[MeSH Terms] OR "Malawi"[All Fields] OR "malawi s"[All Fields]) OR ("Nigeria"[MeSH Terms] OR "Nigeria"[All Fields] OR "nigeria s"[All Fields]) OR "Uganda"[All Fields] OR "uganda s"[All Fields]) OR "Burkina Faso"[Title/Abstract] OR "Cote d'Ivoire"[Title/Abstract] OR "Kenya"[Title/Abstract] OR "Malawi"[Title/Abstract] OR "Nigeria"[Title/Abstract] OR "Uganda"[Title/Abstract] |
| #3 | (("Antimalarials"[Mesh]) OR "Antimalarials" [Pharmacological Action] OR "Antimalarials"[nm] OR anti-malarial [Title/Abstract] OR ("Amodiaquine"[nm] OR "Artesunate"[Mesh] OR "Amodiaquine"[Title/Abstract]) OR "amodiaquine, artesunate drug combination"[nm] OR ("Lumefantrine"[Mesh] OR "Lumefantrine"[Title/Abstract] "Artemether, Lumefantrine Drug Combination"[nm]) OR ("Pyrimethamine"[nm] OR "Pyrimethamine"[Mesh] OR "Pyrimethamine"[Title/Abstract]) OR ("Sulfadoxine"[Mesh] OR "Sulfadoxine"[nm] OR "Sulfadoxine"[Title/Abstract]) OR "fanasil, pyrimethamine drug combination"[nm] OR ("Chloroquine"[Mesh] OR "Chloroquine"[Title/Abstract]) OR "Artesunate"[Mesh] OR ("Quinine"[Mesh] OR "Quinine"[Title/Abstract]) OR (amodiaquine OR atovaquone OR artemisinin OR arteether OR artesunate OR artemether OR artemotil OR azithromycin OR artekin OR chloroquine OR chlorproguanil OR cycloguanil OR clindamycin OR coartem OR dapsone OR desethylamodiaquine OR dihydroartemisinin OR duo-cotecxin OR doxycycline OR halofantrine OR lumefantrine OR lariam OR malarone OR mefloquine OR naphthoquine OR naphthoquinone OR piperaquine OR primaquine OR proguanil OR pyrimethamine OR pyronaridine OR quinidine OR quinine OR riamet OR sulphadoxine OR tetracycline OR tafenoquine)))) |
| #4 | ("Mdr1 protein, Plasmodium falciparum"[nm] OR "pfmdr"[Title/Abstract] OR "pfmdr1"[Title/Abstract] OR "pfmdr"[Title/Abstract] OR "mdr"[Title/Abstract] OR "mdr1"[Title/Abstract] OR "mdr-1"[Title/Abstract] OR "plasmodium falciparum multidrug resistance"[Title/Abstract] OR "plasmodium falciparum multi-drug resistance"[Title/Abstract] OR "plasmodium falciparum multidrug resistant"[Title/Abstract] OR "plasmodium falciparum multi-drug resistant"[Title/Abstract] OR "PfCRT protein, Plasmodium falciparum"[nm] OR "pfcrt"[Title/Abstract] OR "crt"[Title/Abstract] OR "plasmodium falciparum chloroquine resistance transporter"[Title/Abstract] OR "Multidrug Resistance-Associated Proteins"[nm] OR "Tetrahydrofolate Dehydrogenase"[nm] OR "Dihydropteroate Synthase"[nm] OR "PfDHFR"[Title/Abstract] OR "DHFR protein, Plasmodium falciparum"[nm] OR "PfDHPS"[Title/Abstract] OR Pfmdr1 OR Pfmdr-1 OR pfmdr OR mdr OR mdr1 OR mdr-1 OR plasmodium falciparum multidrug resistance OR plasmodium falciparum multi-drug resistance OR plasmodium falciparum multidrug resistant OR plasmodium falciparum multi-drug resistant OR Pfcrt OR crt OR plasmodium falciparum chloroquine resistance transporter OR Pfdhfr) |
| #5 | #1 AND #2 AND #3 AND #4 |

*Table S2. Selection criteria for identified studies*

| Inclusion criteria | |
| --- | --- |
| Participants infected with *P. falciparum* only | Prevalence data of one or more of the selected molecular mutations |
| Samples were collected or originated in Burkina Faso, Cote d’Ivoire, Nigeria, Kenya, Malawi or Uganda | Sample collection occurred between 2000 and 2021, with details of collection year |
| All study designs | Collected blood sample genotyped before treatment |
| Exclusion criteria | |
| Infection with non-*falciparum* malaria or mixed infection | Treatment was received within last month (or date not specified) before genotyping or only treatment failure genotyped |
| Studies with samples collected before 2000, after 2021 or collection year not given | Presence of severe malaria |
| Infection occurred in country other than those specified, or data pooled according to region | Use of laboratory strains, laboratory culture or membrane feeding assays |
| Studies that have grouped multiple years together (>3 years), with no separation of years | Pooled SNP data |
| Number of samples genotyped is not given or is unclear | Prevalence data of treated and untreated samples pooled |
| Original samples used in multiple publications | Use of new or not validated assay |
| Year of sample collection not given | Publication not accessible through UniBas or Swiss TPH servers |
| Publication in a language other than English | Review articles, conference abstract/poster, opinion/commentary, letter or protocol |
