## Supplementary material 2 for "Changes in the prevalence of antimalarial partner drug resistance markers and policy in six sub-Saharan African countries from 2000 to 2021: A systematic review"

**Supplementary information 2**

Table S1. Characteristics of the included studies by region and country, including the author, year(s) of collection and the antimalarial resistance markers that were analysed.

| Region and Country | Author | Year of collection | Markers collected |
| --- | --- | --- | --- |
| East Africa |  |  |  |
| *Kenya*  *(n = 28)* | Akala HM et al. (1) | 2007-2008 | *Pfmdr1* |
|  | Akala HM et al. (2) | 2008-2012 | *Pfcrt, Pfmdr1* |
|  | Bonizzoni M et al. (3) | 2006 | *Pfcrt, Pfmdr1, Pfdhfr, Pfdhps* |
|  | Chebore W et al. (4) | 2016-2017 | *Pfcrt, Pfmdr1* |
|  | Cheruiyot J et al. (5) | 2010-2011 | *Pfcrt, Pfmdr1* |
|  | Eyase FL et al. (6) | 2008-2011 | *Pfcrt, Pfmdr1* |
|  | Hemming-Schroeder E et al. (7) | 2003, 2005, 2008, 2015 | *Pfcrt, Pfmdr1, Pfdhfr, Pfdhps* |
|  | Holmgreen G et al. (8) | 2003 | *Pfcrt, Pfmdr1* |
|  | Juma DW et al. (9) | 2008-2012 | *Pfdhfr, Pfdhps* |
|  | Kiarie WC et al. (10) | 2013 | *Pfcrt* |
|  | Lucchi NW et al (11) | 2010-2013 | *Pfdhfr, Pfdhps* |
|  | Lucchi NW et al. (A) (12) | 2010-2013 | *Pfcrt, Pfmdr1* |
|  | Mang’era CM et al. (13) | 2008 | *Pfcrt, Pfmdr1* |
|  | Oesterholt MJAM et al. (14) | 2003-2004 | *Pfdhfr* |
|  | Muiruri P et al. (15) | 2012-2013 | *Pfmdr1* |
|  | Musyoka KB et al. (16) | 2015 | *Pfcrt, Pfmdr1* |
|  | Mwai L et al. (17) | 2000-2003, 2006 | *Pfcrt, Pfmdr1, Pfdhfr, Pfdhps* |
|  | Okombo J et al. (18) | 2006, 2013 | *Pfcrt, Pfmdr1, Pfdhfr* |
|  | Ostoi V et al. (19) | 2019 | *Pfcrt, Pfmdr1, Pfdhfr, Pfdhps* |
|  | Pacheco MA et al. (20) | 2005, 2010, 2017-2018 | *Pfdhfr, Pfdhps* |
|  | Spalding MD et al. (21) | 2003-2005 | *Pfcrt, Pfmdr1, Pfdhfr, Pfdhps* |
|  | Thwing JI et al. (22) | 2007 | *Pfcrt, Pfmdr1* |
|  | Torrevillas BK et al. (23) | 2015-2018 | *Pfdhfr, Pfdhps* |
|  | Wamae K et al. (24) | 2015-2018 | *Pfcrt, Pfmdr1, Pfdhps* |
|  | Wamae K et al. (25) | 2007-2011, 2014 | *Pfmdr1* |
|  | Wakoli DM et al. (26) | 2018-2021 | *Pfcrt, Pfmdr1, Pfdhfr, Pfdhps* |
|  | Wamae K & Ndwiga L et al. (27) | 2018-2019 | *Pfmdr1* |
|  | Zhou Z et al. (28) | 2012, 2017 | *Pfcrt, Pfmdr1, Pfdhfr, Pfdhps* |
| *Malawi*  *(n = 14)* | Artimovich et al. (29) | 2007-2009, 2012 | *Pfdhfr, Pfdhps* |
|  | Bell et al. (30) | 2003-2005 | *Pfcrt, Pfmdr1, Pfdhfr, Pfdhps* |
|  | Bridges et al. (31) | 2007 | *Pfcrt, Pfdhfr, Pfdhps* |
|  | Bwijo et al. (32) | 2000 | *Pfdhfr, Pfdhps* |
|  | Dzinjalamala et al. (33) | 2000 | *Pfdhfr, Pfdhps* |
|  | Frosch AEP et al. (34) | 2009 | *Pfcrt* |
|  | Gutman J et al. (35) | 2010-2011 | *Pfdhfr, Pfdhps* |
|  | Gutman J et al. (A) (36) | 2009-2011 | *Pfdhps* |
|  | Kublin JG et al. (37) | 2000-2001 | *Pfcrt, Pfmdr1, Pfdhfr* |
|  | Laufer MK et al. (38) | 2005 | *Pfcrt* |
|  | Laufer MK et al. (39) | 2007-2009 | *Pfcrt* |
|  | Lin JT et al. (40) | 2003-2006 | *Pfdhfr, Pfdhps* |
|  | Mita T et al. (41) | 2000 | *Pfcrt, Pfmdr1* |
|  | Mita T et al. (42) | 2000 | *Pfcrt, Pfmdr1* |
| *Uganda*  *(n = 27)* | Asua V et al. (43) | 2016-2017 | *Pfcrt, Pfmdr1, Pfdhfr, Pfdhps* |
|  | Balikagala B et al. (44) | 2013-2018 | *Pfcrt, Pfmdr1* |
|  | Conrad MD et al (A) (45) | 2014 | *Pfmdr1, Pfdhfr, Pfdhps* |
|  | Cuu G et al. (46) | 2015-2016 | *Pfcrt, Pfmdr1* |
|  | Dokomajilar C et al. (47) | 2004-2005 | *Pfmdr1* |
|  | Francis D et al. (48) | 2002-2004 | *Pfcrt, Pfdhfr, Pfdhps* |
|  | Gasasira AF et al. (49) | 2004-2005 | *Pfdhfr, Pfdhps* |
|  | Kamugisha E et al. (50) | 2009-2010 | *Pfcrt, Pfmdr1* |
|  | Kassaza K et al. (51) | 2010, 2015 | *Pfcrt* |
|  | Kiwuwa MS et al. (52) | 2007-2008 | *Pfcrt, Pfmdr1* |
|  | Kyabayinze D et al. (53) | 2000-2001 | *Pfdhfr, Pfdhps* |
|  | Lynch C et al. (54) | 2005 | *Pfdhfr, Pfdhps* |
|  | Lynch CA et al. (55) | 2007 | *Pfdhfr* |
|  | Malamba S et al. (56) | 2003-2006 | *Pfdhfr, Pfdhps* |
|  | Manirakiza G et al. (57) | 2020 | *Pfcrt* |
|  | Mbogo GW et al. (58) | 2005, 2007-2012 | *Pfcrt, Pfmdr1, Pfdhfr, Pfdhps* |
|  | Nanakabirwa JI et al. (59) | 2011-2012 | *Pfcrt, Pfmdr1* |
|  | Nayebare P et al. (60) | 2016-2017 | *Pfcrt, Pfmdr1, Pfdhfr, Pfdhps* |
|  | Tukwasibwe S et al. (61) | 2010-2011 | *Pfcrt, Pfmdr1* |
|  | Tumwebaze P et al. (62) | 2012-2013, 2015 | *Pfcrt, Pfmdr1, Pfdhfr, Pfdhps* |
|  | Tumwebaze P et al. (63) | 2016-2019 | *Pfcrt, Pfmdr1* |
|  | Yeka A et al. (64) | 2013-2014 | *Pfcrt, Pfmdr1* |
|  | Yeka A et al. (65) | 2015-2016 | *Pfcrt, Pfmdr1* |
|  | Rasmussen SA et al. (66) | 2016 | *Pfcrt, Pfmdr1* |
|  | Malamba SS et al. (67) | 2001-2002 | *Pfdhfr, Pfdhps* |
|  | Paganotti GM et al. (68) | 2007 | *Pfmdr1* |
|  | Tumewebaze P et al. (69) | 2021 | *Pfcrt, Pfmdr1, Pfdhfr, Pfdhps* |
| West Africa |  |  |  |
| *Burkina Faso*  *(n = 22)* | Abdel-Aziz et al. (70) | 2000 | *Pfcrt* |
|  | Baraka et al. (71) | 2005-2006 | *Pfcrt, Pfmdr1* |
|  | Cisse M et al. (72) | 2010 | *Pfdhfr, Pfdhps* |
|  | Coulibaly SO et al. (73) | 2010-2011 | *Pfdhfr, Pfdhps* |
|  | Diallo DA et al. (74) | 2002 | *Pfcrt, Pfmdr1, Pfdhfr, Pfdhps* |
|  | Dokomajikar C et al. (75) | 2004 | *Pfcrt, Pfmdr1, Pfdhfr, Pfdhps* |
|  | Gansané A et al. (76) | 2017-2018 | *Pfmdr1* |
|  | Geiger C et al. (77) | 2000, 2009-2012 | *Pfdhfr, Pfdhps* |
|  | Meissner PE et al. (78) | 2003 | *Pfcrt* |
|  | Natama HM et al. (79) | 2013-2016 | *Pfmdr1* |
|  | Oster N et al. (80) | 2001-2002 | *Pfcrt, Pfmdr1* |
|  | Somé AF et al. (81) | 2006 | *Pfcrt, Pfmdr1, Pfdhfr, Pfdhps* |
|  | Somé AF et al. (82) | 2009 | *Pfcrt, Pfmdr1, Pfdhfr, Pfdhps* |
|  | Somé AF et al. (83) | 2012 | *Pfcrt, Pfmdr1, Pfdhfr, Pfdhps* |
|  | Somé AF et al. (84) | 2017 | *Pfcrt, Pfmdr1* |
|  | Sondo P et al. (85) | 2011 | *Pfcrt, Pfmdr1* |
|  | Tinto H et al. (86) | 2001 | *Pfcrt* |
|  | Tinto H et al. (87) | 2003 | *Pfdhfr, Pfdhps* |
|  | Ruizendaal E et al. (88) | 2014-2015 | *Pfdhfr, Pfdhps* |
|  | Zongo I et al. (89) | 2005 | *Pfdhfr, Pfdhps* |
|  | Ehrlich HY et al. (90) | 2019 | *Pfcrt, Pfmdr1* |
|  | Zongo I et al. (91) | 2009-2010 | *Pfcrt, Pfmdr1, Pfdhfr, Pfdhps* |
| *Cote d’Ivoire*  *(n = 7)* | Ako BA et al. (92) | 2005 | *Pfcrt, Pfdhfr, Pfdhps* |
|  | Bla BK et al. (93) | 2006-2007 | *Pfcrt* |
|  | Dagnogo O et al. (94) | 2015 | *Pfcrt* |
|  | Djaman JA et al. (95) | 2001 | *Pfcrt, PFMDR1* |
|  | Djaman JA et al. (A) (96) | 2000-2001 | *Pfdhfr, Pfdhps* |
|  | Konate A et al. (97) | 2013-2014, 2016 | *Pfcrt* |
|  | Ouattara L et al. (98) | 2007 | *Pfcrt, Pfdhfr* |
| *Nigeria*  *(n = 29)* | Adam R et al. (99) | 2018 | *Pfcrt, PFMDR1* |
|  | Agbonlahor DE et al. (100) | 2006 | *Pfmdr1* |
|  | Agomo CO et al. (101) | 2014 | *Pfcrt, PFMDR1* |
|  | Balogun ST et al. (102) | 2010 | *Pfcrt, Pfmdr1* |
|  | Chijioke-Nwauche I et al. (103) | 2010-2011 | *Pfcrt, Pfmdr1, PFDHPS* |
|  | Dokunmu TM et al. (104) | 2018 | *Pfmdr1* |
|  | Emilia AE et al. (105) | 2013 | *Pfmdr1* |
|  | Esu E et al. (106) | 2013-2014 | *Pfdhfr, Pfdhps* |
|  | Fagbemi KA et al. (107) | 2018-2019 | *Pfdhfr, Pfdhps* |
|  | Happi TC et al. (108) | 2003-2004 | *Pfdhfr, Pfdhps* |
|  | Happi TC et al. (109) | 2005 | *Pfcrt, Pfmdr1* |
|  | Happi TC et al. (110) | 2006-2007 | *Pfmdr1* |
|  | Ikegbunam MN et al. (111) | 2014-2015 | *Pfcrt, Pfmdr1* |
|  | Iwalokun BA et al. (112) | 2011 | *Pfdhfr, Pfdhps* |
|  | Kayode AT et al. (113) | 2014-2015 | *Pfcrt, Pfmdr1* |
|  | Kayode AT et al. (114) | 2014-2015 | *Pfdhfr, Pfdhps* |
|  | Oboh MA et al. (115) | 2016-2017 | *Pfcrt, Pfdhfr, Pfdhps* |
|  | Ojurongbe O et al. (116) | 2004-2005 | *Pfcrt, Pfmdr1* |
|  | Ojurongbe O et al. (117) | 2006-2007 | *Pfcrt, Pfmdr1, Pfdhfr* |
|  | Oladipo OO et al. (118) | 2007-2008 | *Pfcrt, Pfmdr1* |
|  | Olasehinde GI et al. (119) | 2017-2018 | *Pfcrt, Pfmdr1* |
|  | Oyebola KM et al. (120) | 2016 | *Pfcrt, Pfmdr1, Pfdhfr, Pfdhps* |
|  | Quan H et al. (121) | 2010-2011, 2013-2014 | *Pfdhfr, Pfdhps* |
|  | Soniran OT et al. (122) | 2013-2014 | *Pfcrt* |
|  | Tola M et al. (123) | 2014 | *Pfcrt, Pfmdr1* |
|  | Wang X et al. (124) | 2016-2020 | *Pfcrt, Pfmdr1, Pfdhfr, Pfdhps* |
|  | Zhao D et al. (125) | 2012-2019 | *Pfcrt, Pfmdr1, Pfdhfr, Pfdhps* |
|  | Olukosi YA et al. (126) | 2000-2002 | *Pfcrt* |
|  | Olukosi YA et al. (127) | 2021 | *Pfdhps* |
| Multi country |  |  |  |
| *(n =11)* | Pearce RJ et al. (128) | 2006 | *Pfdhps* |
|  | Zhou RM et al. (129) | 2012-2015 | *Pfcrt* |
|  | Baraka V et al. (130) | 2012-2014 | *Pfdhps* |
|  | Baraka V et al. (131) | 2012-2014 | *Pfmdr1* |
|  | Wang X et al. (132) | 2016-2018 | *Pfcrt* |
|  | Yan H et al. (133) | 2017-2019 | *Pfdhfr, Pfdhps* |
|  | Beshir KB et al. (134) | 2016 | *Pfcrt, Pfmdr1, Pfdhfr, Pfdhps* |
|  | Foguim ET et al. (135) | 2017-2018 | *Pfcrt* |
|  | Ndong Ngomo JM et al. (136) | 2008-2009 | *Pfcrt* |
|  | Xu C et al. (137) | 2013-2016 | *Pfdhfr, Pfdhps* |
|  | Zhang T et al. (138) | 2012-2016 | *Pfcrt, Pfmdr1* |

*Table S2: Additional characteristics of included studies such as study design, genotyping method and age by country.*

| Study Design | *Cross-Section* | *TES* | *Cohort* | *RCT* | *Not specified* |
| --- | --- | --- | --- | --- | --- |
| *Kenya* | 16 | 6 | 4 | 2 | - |
| *Malawi* | 7 | 7 | - | - | - |
| *Uganda* | 12 | 7 | 4 | 3 | 1 |
| *Burkina Faso* | 8 | 13 | 2 | 1 | - |
| *Cote d’Ivoire* | 4 | 3 | - | - | - |
| *Nigeria* | 20 | 8 | - | 1 | - |
| *Multi-country* | 9 | - | - | 2 | - |
| Genotyping method | ***RFLP*** | ***PCR*** | ***FRET*** | ***LDR*** | ***Other****ª* |
| *Kenya* | 7 | 21 | - | - | 1 |
| *Malawi* | 4 | 7 | - | - | 3 |
| *Uganda* | 10 | 3 | 4 | 11 | 3 |
| *Burkina Faso* | 15 | 4 | - | 1 | 2 |
| *Cote d’Ivoire* | 3 | 4 | - | - | - |
| *Nigeria* | 14 | 10 | 2 | - | 3 |
| *Multi-country* | 4 | 6 | - | - | 1 |
| Age | ***<18*** | ***3-59 months**** | ***>18*** | ***All ages*** | ***Not specified*** |
| *Kenya* | 15 | 2 | 1 | 8 | 4 |
| *Malawi* | 9 | 4 | 4 | 1 | 2 |
| *Uganda* | 14 | 5 | 2 | 9 | 3 |
| *Burkina Faso* | 9 | 7 | 5 | 9 | - |
| *Cote d’Ivoire* | 4 | 3 | - | 2 | 1 |
| *Nigeria* | 10 | 2 | 6 | 12 | 1 |
| *Multi-country* | 4 | 2 | 6 | 1 |  |

*Some studies used more than one Study design/genotyping method/age group. *3-59 months are included in the <18 category too; ª Other may include Enzyme-linked Immunosorbent Assay (ELISA), High Resolution Melt (HRM), Microarray, Molecular Inversion Probe (MIP) or Pyrosequencing. TES: Therapeutic Efficacy Study; RCT: Randomised control trial; RFLP: Restriction Fragment Length Polymorphism; PCR: Polymerase Chain Reaction and sequencing; FRET: Fluorescence Resonance Energy Transfer; LDR: Ligase Detection Reaction.*

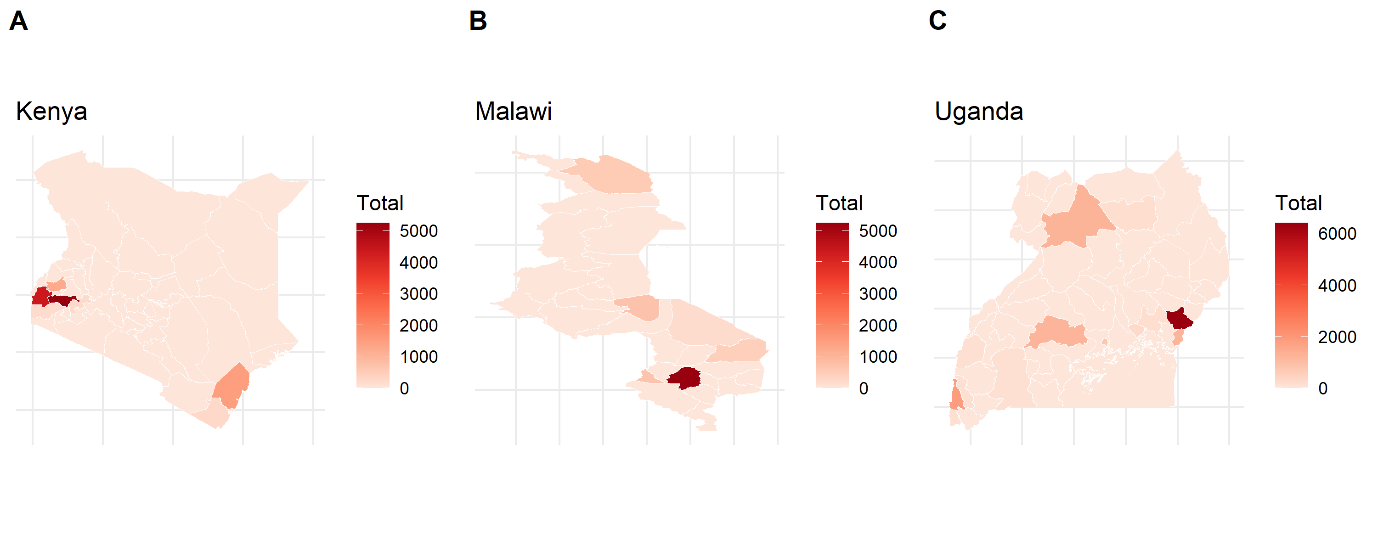

*Figure S1: Number of samples genotyped in each country in East Africa by administrative district. Kenya: 47 districts; Malawi: 28 districts; Uganda: 58 districts. Note: Samples collected across multiple districts not included. Samples genotyped for multiple genes were only included once. Total scales are not the same for each country.*

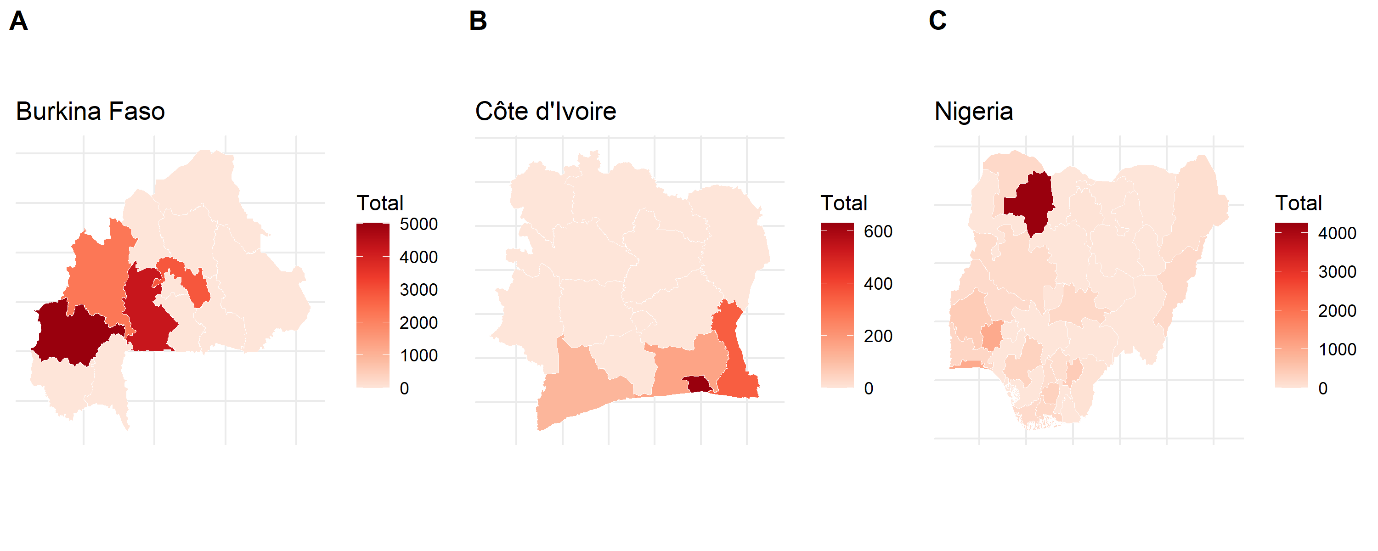

*Figure S2: Number of samples genotyped in each country in West Africa by administrative district; Burkina Faso: 13 districts; Cote d’Ivoire: 14 districts; Nigeria: 37 districts. Note: Samples collected across multiple districts not included. Samples genotyped for multiple genes were only included once. Total scales are not the same for each country.*

***Pfcrt* 76T mutation estimated prevalence**
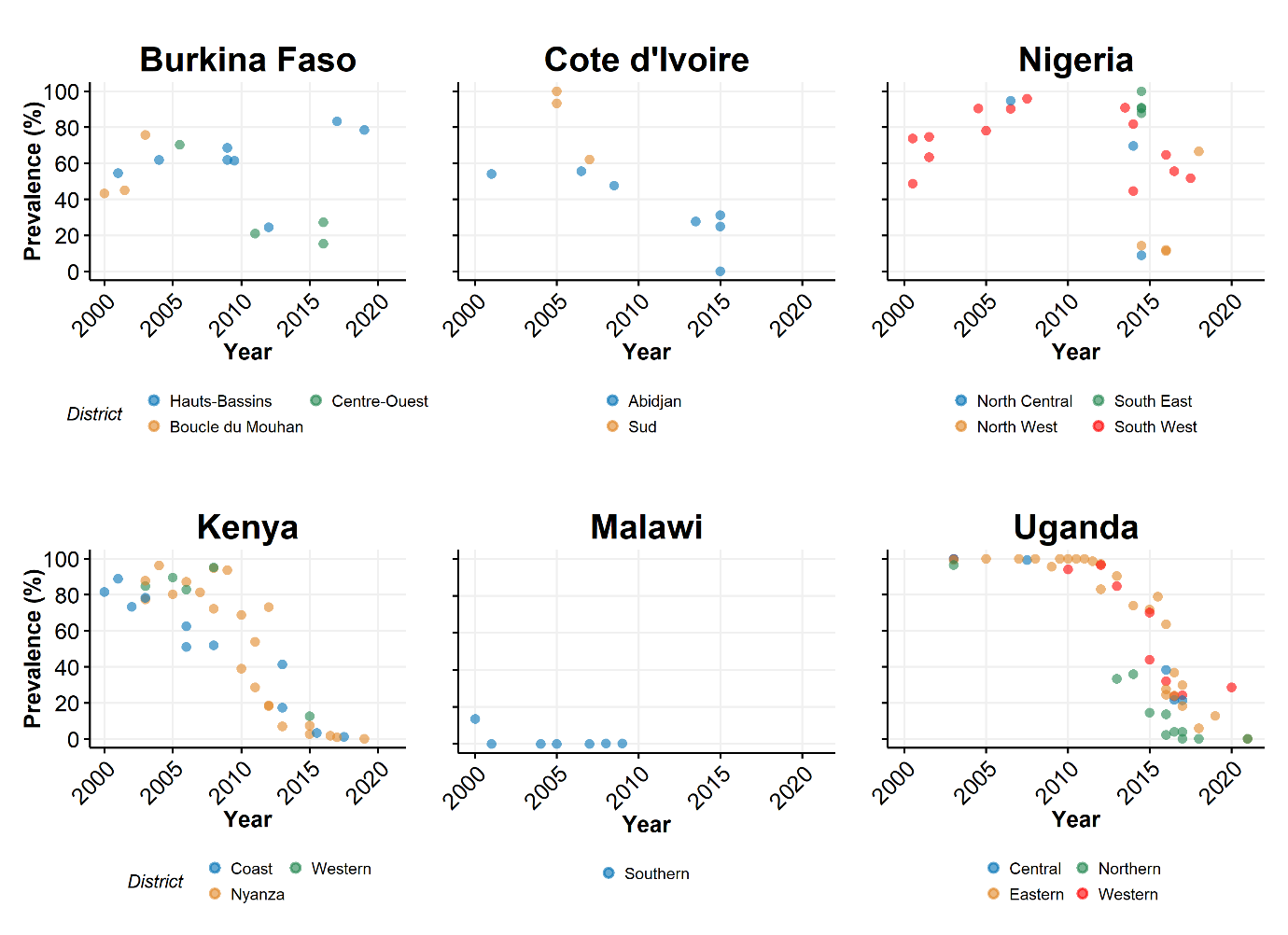

*Figure S3. Prevalence of Pfcrt 76T mutation in each country and the regions within the country that reported mutation data. Districts in some countries were grouped into regions for simplicity. Colours correspond to the region listed. Only regions with greater than or equal to four data points were included to show temporal prevalence.*

***Pfmdr1* mutation estimated prevalence**

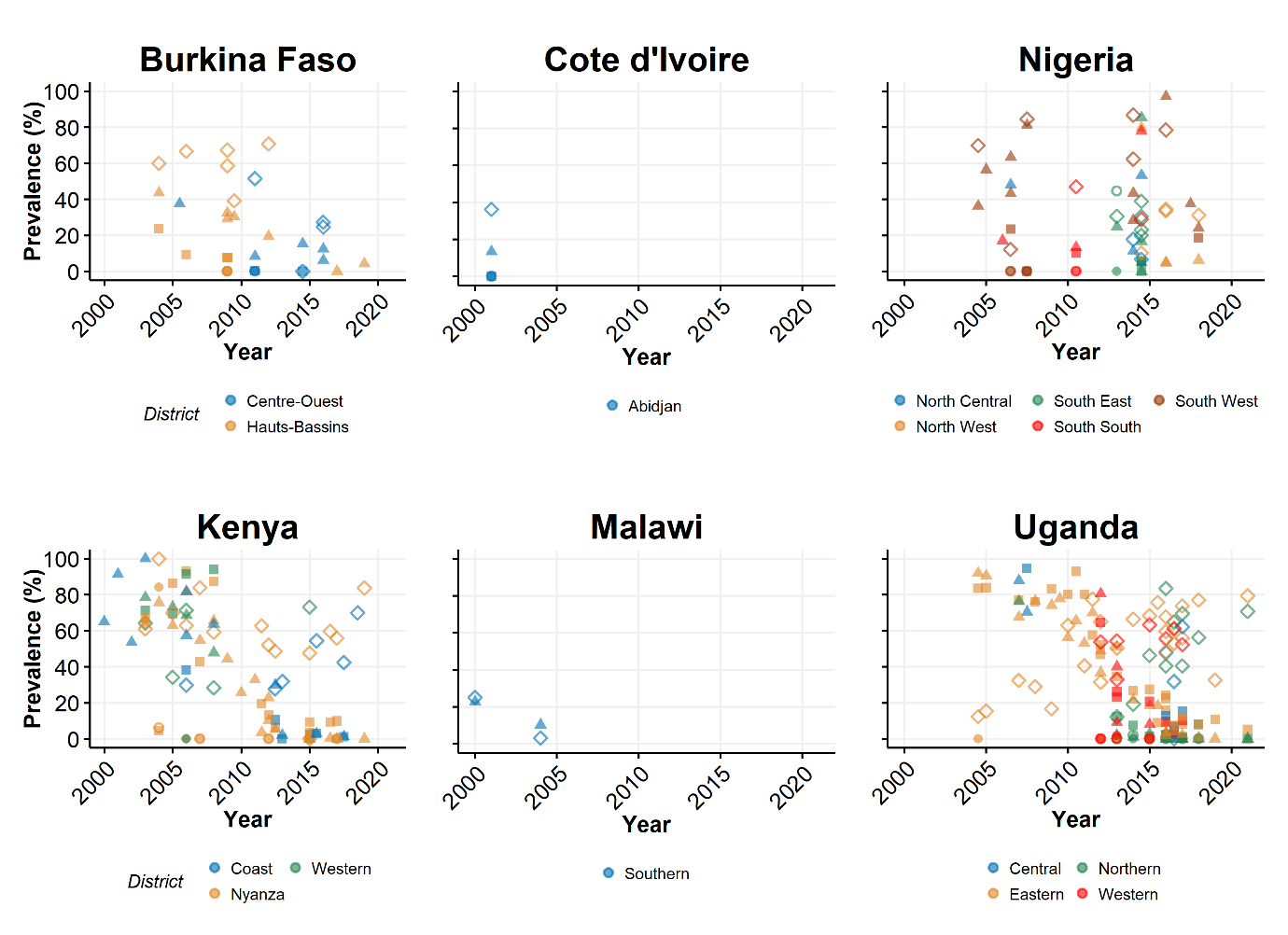

*Figure S4. Prevalence of Pfmdr1 mutations in each country and the regions within the country that reported mutation data. Districts in some countries were grouped into regions for simplicity. Colours correspond to the region listed. Mutations are represented by the following symbols: ▲ N86Y, ◊ Y184F, Օ S1034C, ● N1043D, ■ D1246Y. Only regions with greater than or equal to four data points were included to show temporal prevalence.*

***Pfdhfr* mutation estimated prevalence**

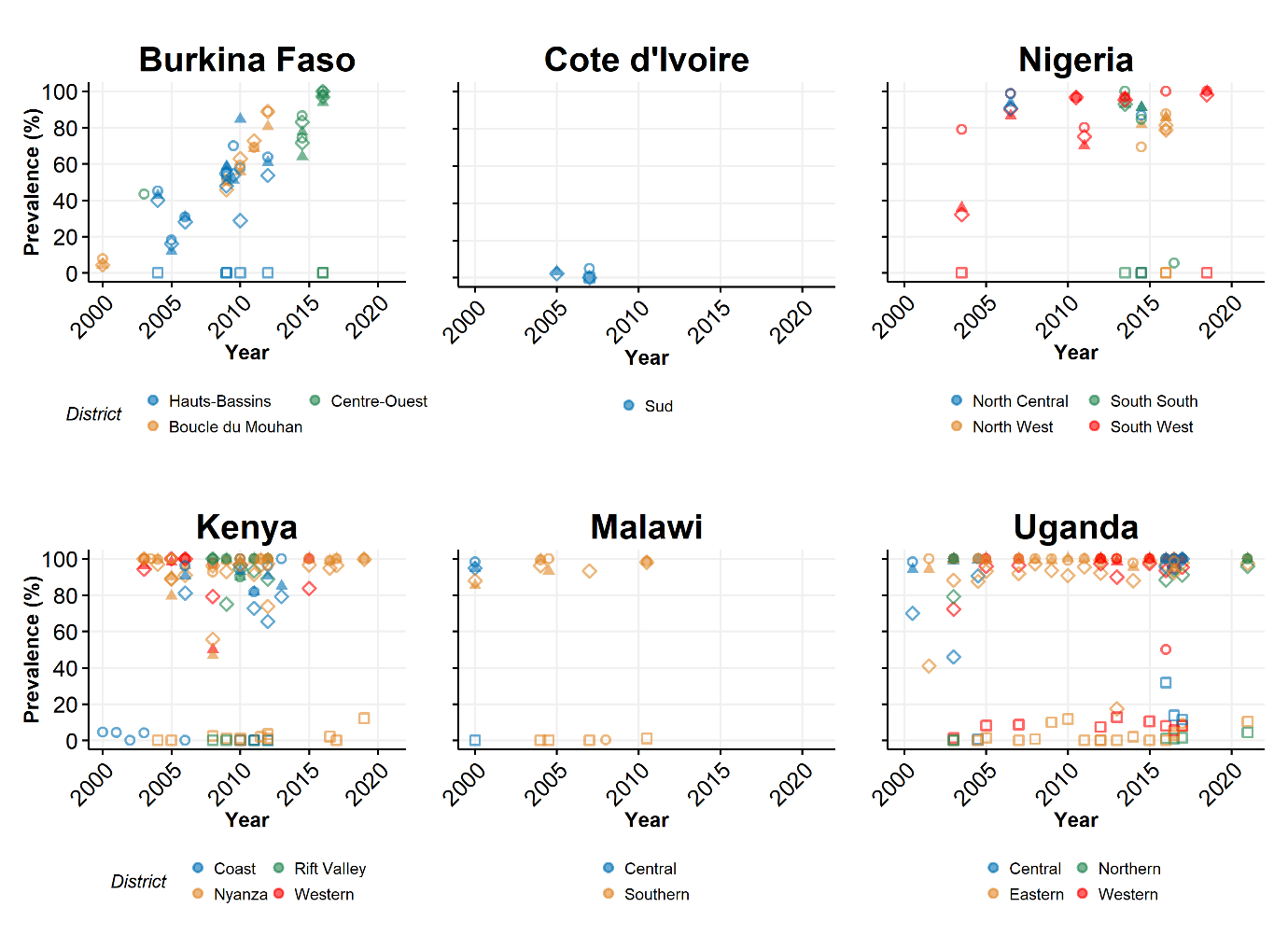

*Figure S5: Prevalence of Pfdhfr mutations in each country and the regions within the country that reported mutation data. Districts in some countries were grouped into regions for simplicity. Colours correspond to the region listed. Mutations are represented by the following symbols: ▲ N51I, ◊ C59R, Օ S108N, □ I164L. Only regions with greater than or equal to four data points were included to show temporal prevalence.*

***Pfdhps* mutation estimated prevalence**

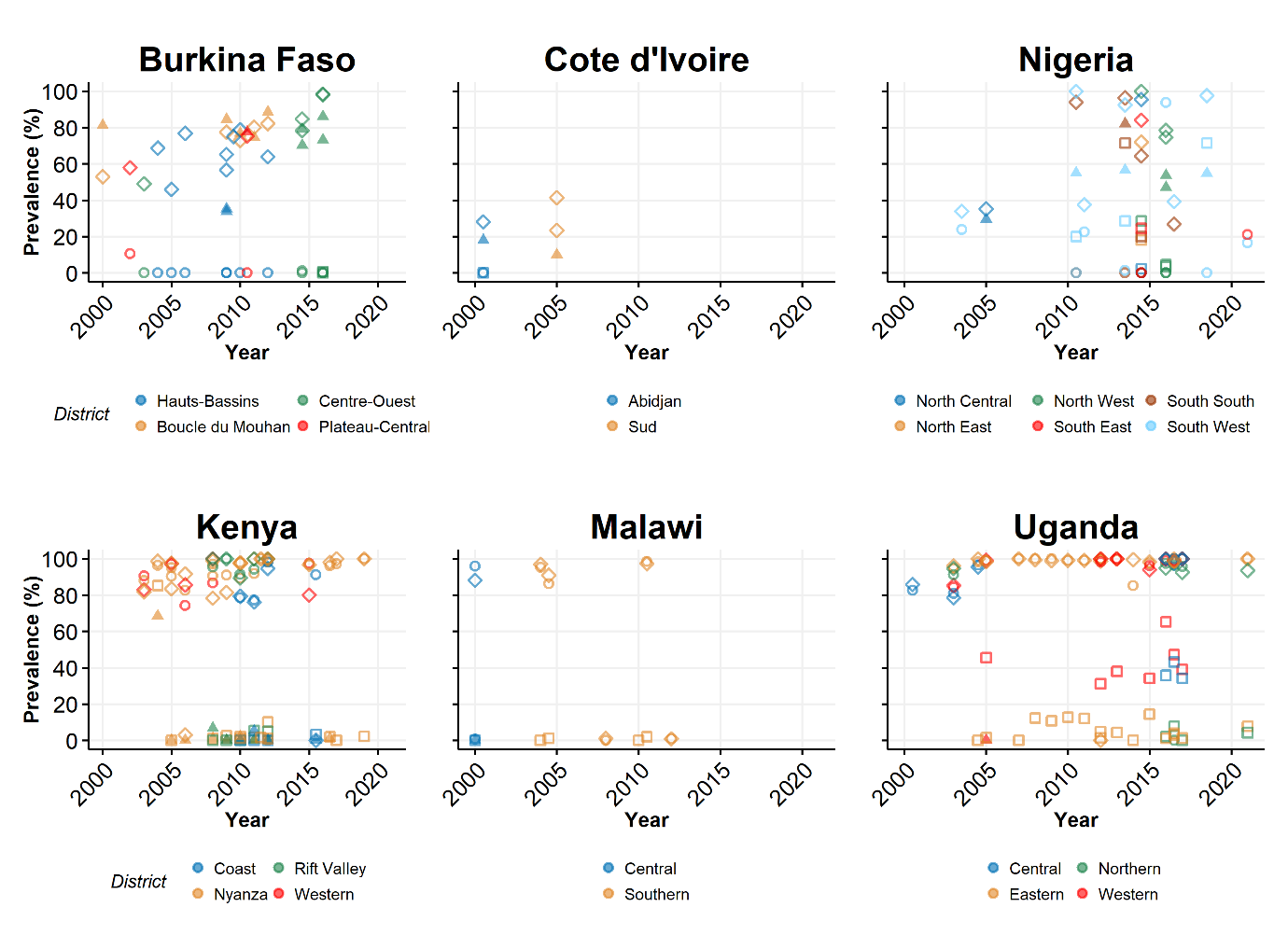

*Figure S6: Prevalence of Pfdhps mutations in each country and the regions within the country that reported mutation data. Districts in some countries were grouped into regions for simplicity. Colours correspond to the region listed. Mutations are represented by the following symbols: ▲ S436A, ◊ A437G, Օ K540E, □ A581G. Only regions with greater than or equal to four data points were included to show temporal prevalence.*

**References included in Systematic Review**

98.	Ouattara L, Bla K, Assi S, Yavo W, Djaman A. PFCRT and DHFR-TS Sequences for Monitoring Drug Resistance in Adzopé Area of Côte d'Ivoire After the Withdrawal of Chloroquine and Pyrimethamine. Tropical Journal of Pharmaceutical Research. 2011;9.
